# Identification of Causal Risk Factors for Pan-Cancers: a Mendelian Randomization Study

**DOI:** 10.1101/2024.07.06.24309341

**Authors:** Bowen Du, Li Fan, Chaopeng Tang, Song Xu, Jingping Ge, Xuejun Shang

**Affiliations:** Department of Urology, Jinling Hospital, Affiliated Hospital of Medical School, Nanjing University, Nanjing, China

**Keywords:** Plasma protein, Plasma metabolites, Cancer, Mendelian randomization, SNP

## Abstract

**Background:** Evidence from observational studies and clinical trials suggests an association between plasma protein and metabolite levels and cancers. However, the causal relationship between them is still unclear.

**Methods:** We collected genome-wide association study (GWAS) summary statistics of plasma protein levels from the UK Biobank Pharma Proteomics Project (UKB-PPP, 9,216 to 34,090 participants) and plasma metabolites from the GWAS Catalog (3,441 to 8,299 participants), paired with summary statistics of 99 types of cancers from FinnGen database (131,348 to 412,181 participants). We conducted univariable and multivariable Mendelian randomization (MR) analyses to explore the causal association between plasma protein and metabolites and cancers.

**Results:** We identified 175 plasma proteins and 28 metabolites causally associated with cancers (p < 1 × 10^−5^). Notably, BTN2A1 is causally associated with an increased risk of bone and articular cartilage cancer (OR = 1.776, 95% CI = 1.429 - 2.207), colorectal cancer (OR = 1.200, 95% CI = 1.129 - 1.275), eye and adnexa cancer (OR = 2.686, 95% CI = 1.943 - 3.714), lip cancer (OR = 3.004, 95% CI = 2.193 - 4.114), oral cancer (OR = 1.905, 95% CI = 1.577 - 2.302), ovary cancer (OR = 1.265, 95% CI = 1.143 - 1.400), and rectum cancer (OR = 1.393, 95% CI = 1.263 - 1.536). N6- carbamoylthreonyladenosine level is causally associated with various cancers including colorectal cancer (OR = 1.800, 95% CI = 1.444 – 2.243), head and neck cancer (OR = 2.423, 95% CI = 1.665 – 3.525), hepatocellular carcinoma (OR = 6.476, 95% CI = 2.841 – 14.762), oral cancer and skin cancer (OR = 1.271, 95% CI = 1.161 – 1.392). Additionally, all results are available at the online database (www.causal-risk.net).

**Conclusions:** Our MR analysis reveals causal risk factors for cancers.

## Introduction

Cancer represents a significant public health challenge globally ^1^. Genome-wide association studies (GWAS) have successfully identified numerous cancer susceptibility loci. However, the underlying mechanisms of most loci and specific gene variants remain unclear, which hampers the translation of genetic susceptibility loci into novel therapeutic targets. Mendelian randomization (MR) provides an approach to understanding the signaling pathways in cancer risk and development by elucidating causal relationships between risk factors and the disease using the analysis of genetic variants in observational data.

Previous studies identified several plasma proteins and metabolites as the risk factors of various cancers ^2^. However, the causality underlying these factors has not been fully explored, primarily due to unmeasured confounding variables or reverse causation. Advances in large-scale GWAS on circulating proteins and metabolites and the methodology of MR have enabled researchers to investigate these causal relationships. Prior investigations have demonstrated the causal effects of certain plasma proteins and metabolites on cancer risk. Nevertheless, comprehensive investigations into the causal effects of plasma proteins and metabolites across a broader range of cancers remain limited.

In this study, we conducted a comprehensive two-sample univariable MR (UVMR) analysis to explore the causal effects of 2,714 plasma proteins, 731 metabolite levels, and 235 metabolite ratios on the risk of 99 types of cancers. To ensure the reliability of our findings, we also performed a multivariable MR (MVMR) analysis. Furthermore, we developed a user-friendly online database, CAURIS (www.causal-risk.net), to help users access the results.

## Methods

### Datasets

All analyses in this investigation relied on summary-level GWAS data. GWAS summary statistics for plasma proteins and metabolites were used as exposures. The GWAS summary statistics for plasma proteins were derived from the UK Biobank Pharma Proteomics Project (UKB-PPP), a large-scale GWAS repository composed of 2,940 proteins. The GWAS summary statistics for plasma metabolites were obtained from a study conducted by Chen et al., which included 1,091 plasma metabolite levels and 309 metabolite ratios.

We employed GWAS summary statistics encompassing 99 types of cancers, sourced from the FinnGen dataset ^3^, which included approximately 370,000 participants, as our outcome data. The cancer types were selected according to the third revision of the International Classification of Diseases for Oncology (ICD-O-3) codes. The description of the endpoints (DF12) can be accessed from the FinnGen website. The FinnGen GWAS summary statistics (release v10) were downloaded from the FinnGen website. The GWAS summary statistics for smoking, alcohol consumption, body mass index (BMI), physical exercise, and chemical exposure were used in MVMR. All the GWAS data used in this study were restricted to European ancestry. Comprehensive details of all GWAS summaries are provided in Table S1 - S4.

### Instrumental variable selection

The genetic instrument variables (IVs), typically single-nucleotide polymorphisms (SNPs), were derived from GWAS summary statistics using the TwoSampleMR R package. For IV clumping, the p-value threshold was set to 1×10^−6^, and other parameters were set with r^2^ < 0.001 and a clumping distance = 250 kb. Linkage disequilibrium (LD) r2 and clumping window were estimated based on 1000 Genomes LD reference panel (European population). During the data harmonization process, both palindromic and duplicated SNPs were excluded.

Instrumental variable (IV) outliers were identified using the MR-PRESSO R package. For each SNP, the MR-PRESSO outlier test calculated a p-value to evaluate its significance for pleiotropy, while the MR-PRESSO global test calculated a p-value for the overall presence of horizontal pleiotropy. SNPs with a p-value less than 0.05 were removed if the global p-value was less than 0.05. Subsequently, the global p-value was recalculated to evaluate the extent of pleiotropy.

### UVMR analysis

We conducted a two-sample UVMR analysis to investigate the causal relationship between plasma proteins and metabolites and cancer risks, employing the TwoSampleMR R package. We utilized three established MR methods for MR analysis with multiple IVs: MR-Egger (MRE), weighted median (WM), and inverse variance weighted (IVW). Analyses with fewer than three IVs were excluded, as an inadequate number of IVs would compromise the reliability of the analysis. Furthermore, we established a multiple testing significance threshold: p < 0.05/n, where n is the number of exposures ^4^.

To mitigate biases introduced by weak IVs, we calculated the F-statistic according to the reference ^5^. Initially, we computed the proportion of variance explained by each IV (PVE) using the equation ^6^:

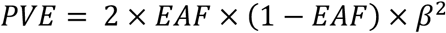

where EAF represents the effect allele frequency (EAF), and β is the effect size. The F-statistic is subsequently calculated using the formulation:

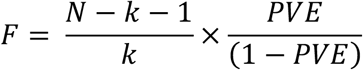

where N is the sample size and k is the number of IVs. We computed the mean F-statistic across all IVs. We used the MR Steiger directionality test to examine whether exposure was directionally causal for the outcome.

### Sensitivity analyses

We evaluated the heterogeneity of MR analysis using Cochran’s Q test, considering a p-value (P_Q_) less than 0.05 as indicative of heterogeneity. Pleiotropy was assessed via MR-Egger regression ^7^ and MR-PRESSO residuals tests, with a p-value (P_Egger_ or P_PRESSO_) less than 0.05 indicating evidence of pleiotropy. P_Q_ and P_Egger_ were calculated using the TwoSampleMR R package, while P_PRESSO_ was computed with the MR-PRESSO R package. To ensure the reliability of our findings, we excluded analyses with P_Q_, P_Egger_, or P_PRESSO_ below 0.05 from the results.

### MVMR analysis

Considering that potential genetic correlations among exposures may introduce inaccuracies in UVMR analyses ^8^, we employed MVMR analysis using the MVMR-IVW method. Since cancer risk is associated with smoking^9^, alcohol consumption ^10^, BMI ^11^, physical exercise ^12^, and chemical exposure ^13^, the analyses were adjusted for these confounding variables within the European population. These MVMR analyses were executed using the TwoSampleMR R package and were conducted separately for each protein and metabolite. The IVs were selected based on the following criteria: a p-value threshold of 1×10^−6^, R^2^ less than 0.001, and a clumping distance of 250 kb.

## Results

### Study design

The design of this study is presented in Figure 1. We utilized UVMR analyses to investigate the causal effect of plasma proteins and metabolites on cancers. GWAS summary statistics of plasma proteins and plasma metabolite were used as exposures, while GWAS summary statistics of cancer risks were used as outcomes. Pleiotropic SNPs identified by the MR-PRESSO outlier test were removed. Exposures with fewer than three IVs were excluded.

**Figure 1.**
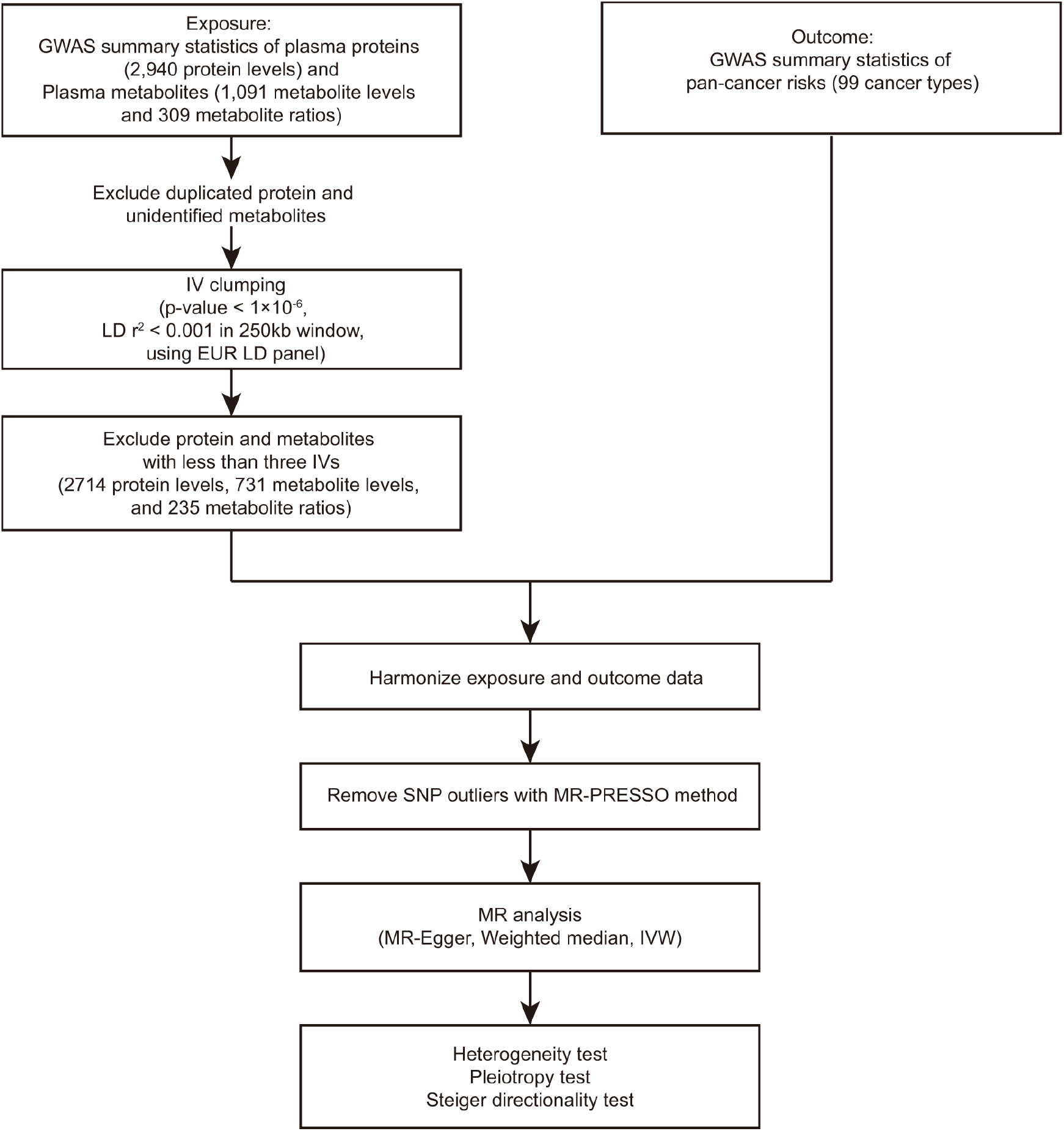
Flowchart of the data collection, processing, and analysis procedures of this study. GWAS summary statistics of plasma proteins and metabolites were used as exposures. GWAS summary statistics of pan-cancer risks were used as outcomes. The instrumental variables (IVs) were extracted and clumped. The IV outliers were removed using the MR-PRESSO method. Mendelian randomization (MR) analyses were conducted using MR-Egger, Weighted median, and IVW methods. The MR Steiger directionality test was used to examine whether the exposure was directionally causal for the outcome. Heterogeneity of the MR analysis was tested using Cochran’s Q test. Pleiotropy of MR analysis was assessed via MR-Egger regression and MR-PRESSO residuals test.

We conducted our MR analysis using MR-Egger, weighted median (WM), and inverse variance weighted (IVW) methods. The heterogeneity of the MR analysis was evaluated with Cochran’s Q test, and the pleiotropy of MR analysis was assessed with MR-Egger regression and MR-PRESSO residuals tests. The significance threshold for the causal associations with plasma proteins and metabolites was set as follows: protein p = 1.84×10^−5^ (0.05/2714), metabolite p = 5.18×10^−5^ (0.05/966). Causal associations with significant heterogeneity or pleiotropy were excluded. We totally identified 694 significant causal associations (p < 1×10^−5^), including 617 causal associations between plasma proteins and cancer risks (Table S5) and 77 causal associations between plasma metabolites and cancer risks (Table S6). All results can be accessed through the online database CAURIS (www.causal-risk.net). We present some of the results in the following context.

### Causal effects of butyrophilins on cancer risks

Butyrophilins (BTN), members of the immunoglobulin superfamily, are associated with the regulation of immune response to tumors. BTN2A1 and BTN3A1 are essential for the activation of human Vγ9Vδ2 T cells. Our study indicated that BTN2A1 is causally associated with an increased risk of bone and articular cartilage cancer (OR = 1.776, 95% CI = 1.429 - 2.207, p = 2.17×10^−7^, IVW), colorectal cancer (OR = 1.200, 95% CI = 1.129 - 1.275, p = 8.55×10^−8^, MR Egger), eye and adnexa cancer (OR = 2.686, 95% CI = 1.943 - 3.714, p = 5.53×10^−8^, MR Egger), lip cancer (OR = 3.004, 95% CI = 2.193 - 4.114, p = 7.18×10^−12^, WM), oral cancer (OR = 1.905, 95% CI = 1.577 - 2.302, p = 2.66×10^−9^, MR Egger), ovary cancer (OR = 1.265, 95% CI = 1.143 - 1.400, p = 5.6×10^−6^, IVW), and rectum cancer (OR = 1.393, 95% CI = 1.263 - 1.536, p = 3.14×10^−9^, MR Egger). We also detected a causal effect of BTN2A1 associated with a reduced risk of clear cell renal cell carcinoma (ccRCC, OR = 0.640, 95% CI = 0.546 - 0.750, p = 3.89×10^−7^, MR Egger), prostate cancer (OR = 0.932, 95% CI = 0.903 - 0.961, p = 6.18×10^−6^, IVW), and urothelial carcinoma (OR = 0.818, 95% CI = 0.767 - 0.873, p = 1.28×10^−9^, IVW).

BTN3A2, another member of the butyrophilins, is associated with mental disorder ^14^ and type 1 diabetes ^15^. Our investigation demonstrated the causal effect of BTN3A2 on the risks of various cancer types. Specifically, BTN3A2 is causally associated with an increased risk of ccRCC (OR = 1.258, 95% CI =, p = 1.35×10^−6^, WM) and urothelial carcinoma (OR = 1.107, 95% CI = 1.061 - 1.154, p = 2.03×10^−6^, IVW). BTN3A2 is causally associated with a decreased risk of cervix uteri cancer (OR = 0.682, 95% CI = 0.591 - 0.787, p = 1.57×10^−7^, WM), colorectal cancer (OR = 0.937, 95% CI = 0.912 - 0.963, p = 2.28×10^−6^, IVW), eye and adnexa cancer (OR = 0.631, 95% CI = 0.522 - 0.763, p = 1.87×10^−6^, WM), lip cancer (OR = 0.542, 95% CI = 0.441 - 0.666, p = 1.95×10^−7^, MR Egger), non-small cell lung cancer (OR = 0.875, 95% CI = 0.849 - 0.902, p = 4.27×10^−18^, IVW), lung squamous cell carcinoma (OR = 0.778, 95% CI = 0.735 - 0.823, p = 1.63×10^−18^, IVW), oral cancer (OR = 0.733, 95% CI = 0.662 - 0.811, p = 1.76×10^−9^, WM), and rectal cancer (OR = 0.864, 95% CI = 0.813 - 0.917, p = 1.8×10^−6^, WM). Thus, our investigation indicates that BTN2A1 and BTN3A2 may participate in cancer development.

### Causal effects of plasma protein on risks of basal cell carcinoma

Basal cell carcinoma (BBC) is the most common malignant tumor in white populations ^16^; however, its pathological mechanisms are not well studied. We identified multiple plasma proteins associated with an increased risk of basal cell carcinoma (Table 2), including ABHD14B (OR = 1.106, 95% CI = 1.063 - 1.150, p = 5.35×10^−7^, IVW), ADAM9 (OR = 1.184, 95% CI = 1.108 - 1.265, p = 5.11×10^−7^, IVW), B2M (OR = 1.207, 95% CI = 1.151 - 1.266, p = 9.0×10^−15^, IVW), EPO (OR = 1.236, 95% CI = 1.147 - 1.332, p = 2.88×10^−8^, IVW), IL1A (OR = 1.806, 95% CI = 1.551 - 2.102, p = 2.46×10^−14^, WM), PDIA5 (OR = 1.118, 95% CI = 1.067 - 1.173, p = 3.34×10^−6^, IVW) and TNFRSF9 (OR = 1.262, 95% CI = 1.183 - 1.347, p = 2.11×10^−12^, IVW). We also identified multiple plasma proteins associated with a decreased risk of basal cell carcinoma, including CSNK1D (OR = 0.740, 95% CI = 0.657 - 0.833, p = 6.53×10^−7^, IVW), F13B (OR = 0.924, 95% CI = 0.893 - 0.955, p = 2.7×10^−6^, IVW), PROCR (OR = 0.906, 95% CI = 0.875 - 0.937, p = 5.63×10^−6^, MR Egger), and SV2A (OR = 0.645, 95% CI = 0.559 - 0.744, p = 1.75×10^−9^, WM). These results may provide insight into the pathological mechanisms by revealing the causal genes associated with cancer risks.

**Table 1.**
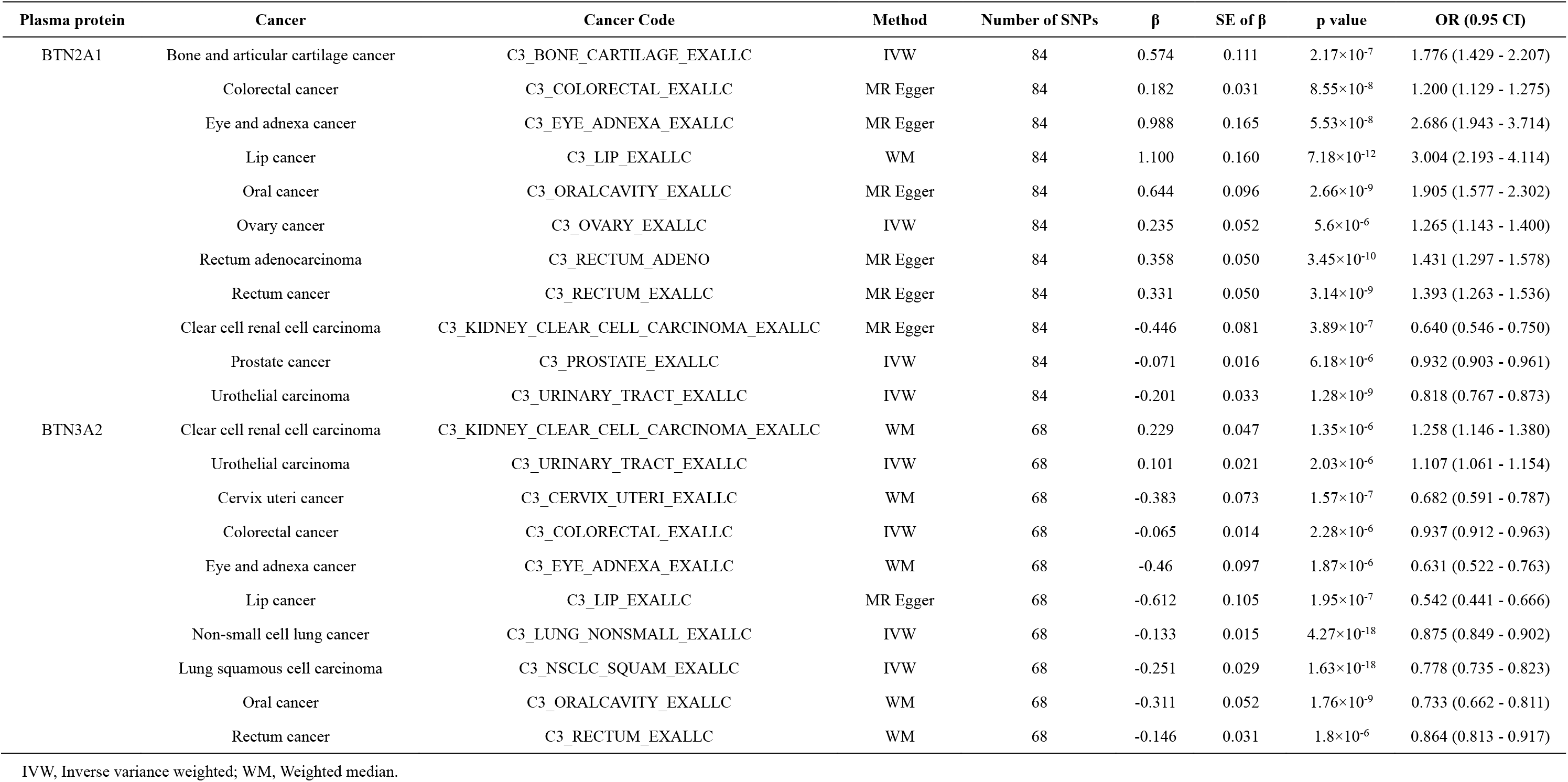
Causal effect of BTN2A1 and BTN3A2 on cancer risks.

| Plasma protein | Cancer | Cancer Code | Method | Number of SNPs | $\beta$ | SE of $\beta$ | p value | OR (0.95 CI) |
| --- | --- | --- | --- | --- | --- | --- | --- | --- |
| BTN2A1 | Bone and articular cartilage cancer | C3_BONE_CARTILAGE_EXALLC | IVW | 84 | 0.574 | 0.111 | $2.17 \times 10^{-7}$ | 1.776 (1.429 - 2.207) |
| | Colorectal cancer | C3_COLORECTAL_EXALLC | MR Egger | 84 | 0.182 | 0.031 | $8.55 \times 10^{-8}$ | 1.200 (1.129 - 1.275) |
| | Eye and adnexa cancer | C3_EYE_ADNEXA_EXALLC | MR Egger | 84 | 0.988 | 0.165 | $5.53 \times 10^{-8}$ | 2.686 (1.943 - 3.714) |
| | Lip cancer | C3_LIP_EXALLC | WM | 84 | 1.100 | 0.160 | $7.18 \times 10^{-12}$ | 3.004 (2.193 - 4.114) |
| | Oral cancer | C3_ORALCAVITY_EXALLC | MR Egger | 84 | 0.644 | 0.096 | $2.66 \times 10^{-9}$ | 1.905 (1.577 - 2.302) |
| | Ovary cancer | C3_OVARY_EXALLC | IVW | 84 | 0.235 | 0.052 | $5.6 \times 10^{-6}$ | 1.265 (1.143 - 1.400) |
| | Rectum adenocarcinoma | C3_RECTUM_ADENO | MR Egger | 84 | 0.358 | 0.050 | $3.45 \times 10^{-10}$ | 1.431 (1.297 - 1.578) |
| | Rectum cancer | C3_RECTUM_EXALLC | MR Egger | 84 | 0.331 | 0.050 | $3.14 \times 10^{-9}$ | 1.393 (1.263 - 1.536) |
| | Clear cell renal cell carcinoma | C3_KIDNEY_CLEAR_CELL_CARCCINOMA_EXALLC | MR Egger | 84 | -0.446 | 0.081 | $3.89 \times 10^{-7}$ | 0.640 (0.546 - 0.750) |
| | Prostate cancer | C3_PROSTATE_EXALLC | IVW | 84 | -0.071 | 0.016 | $6.18 \times 10^{-6}$ | 0.932 (0.903 - 0.961) |
| | Urothelial carcinoma | C3_URINARY_TRACT_EXALLC | IVW | 84 | -0.201 | 0.033 | $1.28 \times 10^{-9}$ | 0.818 (0.767 - 0.873) |
| BTN3A2 | Clear cell renal cell carcinoma | C3_KIDNEY_CLEAR_CELL_CARCCINOMA_EXALLC | WM | 68 | 0.229 | 0.047 | $1.35 \times 10^{-6}$ | 1.258 (1.146 - 1.380) |
| | Urothelial carcinoma | C3_URINARY_TRACT_EXALLC | IVW | 68 | 0.101 | 0.021 | $2.03 \times 10^{-6}$ | 1.107 (1.061 - 1.154) |
| | Cervix uteri cancer | C3_CERVIX_UTERI_EXALLC | WM | 68 | -0.383 | 0.073 | $1.57 \times 10^{-7}$ | 0.682 (0.591 - 0.787) |
| | Colorectal cancer | C3_COLORECTAL_EXALLC | IVW | 68 | -0.065 | 0.014 | $2.28 \times 10^{-6}$ | 0.937 (0.912 - 0.963) |
| | Eye and adnexa cancer | C3_EYE_ADNEXA_EXALLC | WM | 68 | -0.46 | 0.097 | $1.87 \times 10^{-6}$ | 0.631 (0.522 - 0.763) |
| | Lip cancer | C3_LIP_EXALLC | MR Egger | 68 | -0.612 | 0.105 | $1.95 \times 10^{-7}$ | 0.542 (0.441 - 0.666) |
| | Non-small cell lung cancer | C3_LUNG_NONSMALL_EXALLC | IVW | 68 | -0.133 | 0.015 | $4.27 \times 10^{-18}$ | 0.875 (0.849 - 0.902) |
| | Lung squamous cell carcinoma | C3_NSCLC_SQUAM_EXALLC | IVW | 68 | -0.251 | 0.029 | $1.63 \times 10^{-18}$ | 0.778 (0.735 - 0.823) |
| | Oral cancer | C3_ORALCAVITY_EXALLC | WM | 68 | -0.311 | 0.052 | $1.76 \times 10^{-9}$ | 0.733 (0.662 - 0.811) |
| | Rectum cancer | C3_RECTUM_EXALLC | WM | 68 | -0.146 | 0.031 | $1.8 \times 10^{-6}$ | 0.864 (0.813 - 0.917) |

IVW, Inverse variance weighted; WM, Weighted median.

**Table 2.**
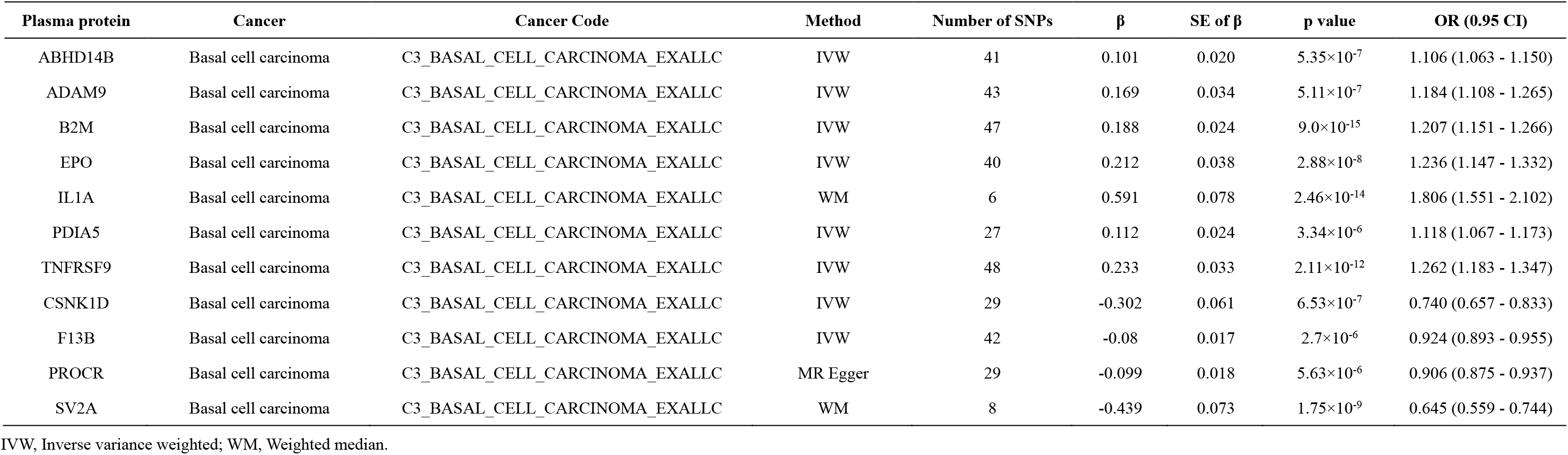
Causal effect of plasma proteins on basal cell carcinoma.

### Causal effects of N6-carbamoylthreonyladenosine on cancer risks

We identified multiple circulating metabolites as factors causally associated with cancer risks (Table 3). For instance, N6-carbamoylthreonyladenosine is causally associated with an increased risk of multiple cancer types, including basal cell carcinoma (OR = 1.299, 95% CI = 1.182 - 1.428, p = 6.31×10^−8^, IVW), cervix uteri cancer (OR = 4.826, 95% CI = 2.408 - 9.672, p = 9.07×10^−6^, IVW), colorectal cancer (OR = 1.800, 95% CI = 1.444 - 2.243, p = 1.63×10^−7^, WM), diffuse large B cell lymphoma (OR = 5.186, 95% CI = 3.055 - 8.804, p = 1.09×10^−9^, WM), eye and adnexa cancer (OR = 8.404, 95% CI = 3.317 - 21.288, p = 7.16×10^−6^, IVW), head and neck cancer (OR = 2.423, 95% CI = 1.665 - 3.525, p = 3.74×10^−6^, WM), hepatocellular carcinoma (OR = 6.476, 95% CI = 2.841 - 14.762, p = 8.86×10^−6^, WM), and non-small cell lung cancer (OR = 1.881, 95% CI = 1.480 - 2.391, p = 2.39×10^−7^, WM).

**Table 3.** Causal effect of N6-carbamoylthreonyladenosine on cancer risks (p < 1×10^−5^).

| Plasma metabolite | Cancer | Cancer Code | Method | Number of SNPs | $\beta$ | SE of $\beta$ | P value | OR (0.95 CI) |
| --- | --- | --- | --- | --- | --- | --- | --- | --- |
| N6-carbamoylthreonyladenosine | Basal cell carcinoma | C3_BASAL_CELL_CARCCINOMA_EXALLC | IVW | 13 | 0.262 | 0.048 | $6.31 \times 10^{-8}$ | 1.299 (1.182 - 1.428) |
| | Cervix uteri cancer | C3_CERVIX_UTERI_EXALLC | IVW | 13 | 1.574 | 0.355 | $9.07 \times 10^{-6}$ | 4.826 (2.408 - 9.672) |
| | Colorectal cancer | C3_COLORECTAL_EXALLC | WM | 13 | 0.588 | 0.112 | $1.63 \times 10^{-7}$ | 1.800 (1.444 - 2.243) |
| | Diffuse large B cell lymphoma | C3_DLBCL_EXALLC | WM | 13 | 1.646 | 0.270 | $1.09 \times 10^{-9}$ | 5.186 (3.055 - 8.804) |
| | Eye and adnexa cancer | C3_EYE_ADNEXA_EXALLC | IVW | 13 | 2.129 | 0.474 | $7.16 \times 10^{-6}$ | 8.404 (3.317 - 21.288) |
| | Head and neck cancer | C3_HEAD_AND_NECK_EXALLC | WM | 13 | 0.885 | 0.191 | $3.74 \times 10^{-6}$ | 2.423 (1.665 - 3.525) |
| | Hepatocellular carcinoma | C3_HEPATOCELLU_CARCCINOMA_EXALLC | WM | 13 | 1.868 | 0.420 | $8.86 \times 10^{-6}$ | 6.476 (2.841 - 14.762) |
| | Non-small cell lung cancer | C3_LUNG_NONSMALL_EXALLC | WM | 13 | 0.632 | 0.122 | $2.39 \times 10^{-7}$ | 1.881 (1.480 - 2.391) |
| | Non-Hodgkin lymphoma | C3_NONHODGKIN_EXALLC | WM | 13 | 1.538 | 0.274 | $2.02 \times 10^{-8}$ | 4.657 (2.721 - 7.972) |
| | Lung squamous cell carcinoma | C3_NSCLC_SQUAM_EXALLC | WM | 13 | 1.198 | 0.241 | $6.49 \times 10^{-7}$ | 3.312 (2.067 - 5.309) |
| | Oral cancer | C3_ORALCAVITY_EXALLC | WM | 13 | 1.302 | 0.311 | $2.93 \times 10^{-5}$ | 3.675 (1.996 - 6.767) |
| | Rectum cancer | C3_RECTUM_EXALLC | WM | 13 | 1.033 | 0.178 | $6.14 \times 10^{-9}$ | 2.810 (1.983 - 3.980) |
| | Skin cancer | C3_SKIN_EXALLC | IVW | 13 | 0.240 | 0.046 | $2.17 \times 10^{-7}$ | 1.271 (1.161 - 1.392) |

IVW, Inverse variance weighted; WM, Weighted median.

### Causal effects of plasma metabolites on risks of breast cancer

Previous studies revealed several plasma metabolites as risk factors for breast cancer ^17,18 19^. In this study, we identified several plasma metabolites causally associated with elevated risks of breast cancer (Table 4), including DHEAS (OR = 1.226, 95% CI = 1.134 - 1.325, p = 2.87×10^−7^, IVW), epiandrosterone sulfate (OR = 1.055, 95% CI = 1.030 - 1.080, p = 8.32×10^−6^, IVW), and androsterone sulfate (OR = 1.052, 95% CI = 1.030 - 1.074, p = 2.69×10^−6^, IVW). We also identified several plasma metabolites associated with reduced risks of breast cancer, including homostachydrine (OR = 0.753, 95% CI = 0.669 - 0.847, p = 2.26×10^−6^, IVW), glycohyocholate (OR = 0.822, 95% CI = 0.745 - 0.907, p = 9.2×10^−5^, IVW), and ribitol (OR = 0.921, 95% CI = 0.889 - 0.954, p = 4.03×10^−6^, IVW).

**Table 4.** Causal effect of plasma metabolites on breast cancers (p < 1×10^−4^).

| Plasma metabolite | Cancer | Cancer Code | Method | Number of SNPs | $\beta$ | SE of $\beta$ | P value | OR (0.95 CI) |
| --- | --- | --- | --- | --- | --- | --- | --- | --- |
| DHEAS | Breast cancer | C3_BREAST_EXALLC | IVW | 9 | 0.204 | 0.040 | $2.87 \times 10^{-7}$ | 1.226 (1.134 - 1.325) |
| Epiandrosterone sulfate | Breast cancer | C3_BREAST_EXALLC | IVW | 14 | 0.053 | 0.012 | $8.32 \times 10^{-6}$ | 1.055 (1.030 - 1.080) |
| Androsterone sulfate | Breast cancer | C3_BREAST_EXALLC | IVW | 21 | 0.050 | 0.011 | $2.69 \times 10^{-6}$ | 1.052 (1.030 - 1.074) |
| Homostachydrine | Breast cancer | C3_BREAST_EXALLC | IVW | 5 | -0.284 | 0.060 | $2.26 \times 10^{-6}$ | 0.753 (0.669 - 0.847) |
| Glycohyocholate | Breast cancer | C3_BREAST_EXALLC | IVW | 7 | -0.196 | 0.050 | $9.2 \times 10^{-5}$ | 0.822 (0.745 - 0.907) |
| Ribitol | Breast cancer | C3_BREAST_EXALLC | IVW | 9 | -0.082 | 0.018 | $4.03 \times 10^{-6}$ | 0.921 (0.889 - 0.954) |
| N-acetylglutamate | HER- breast cancer | C3_BREAST_ERNEG_EXALLC | IVW | 6 | 0.357 | 0.090 | $7.51 \times 10^{-5}$ | 1.429 (1.198 - 1.706) |
| Indolepropionate | HER- breast cancer | C3_BREAST_ERNEG_EXALLC | IVW | 7 | -0.293 | 0.072 | $5.03 \times 10^{-5}$ | 0.746 (0.647 - 0.860) |
| DHEAS | HER+ breast cancer | C3_BREAST_ERPLUS_EXALLC | IVW | 9 | 0.258 | 0.049 | $1.48 \times 10^{-7}$ | 1.294 (1.175 - 1.424) |
| Androsterone sulfate | HER+ breast cancer | C3_BREAST_ERPLUS_EXALLC | IVW | 21 | 0.069 | 0.016 | $1.01 \times 10^{-5}$ | 1.071 (1.039 - 1.105) |
| Ribitol | HER+ breast cancer | C3_BREAST_ERPLUS_EXALLC | IVW | 9 | -0.112 | 0.023 | $1.42 \times 10^{-6}$ | 0.894 (0.854 - 0.936) |
| Homostachydrine | HER+ breast cancer | C3_BREAST_ERPLUS_EXALLC | IVW | 5 | -0.543 | 0.099 | $4.23 \times 10^{-8}$ | 0.581 (0.478 - 0.705) |
| Glyco-beta-muricholate | HER+ breast cancer | C3_BREAST_ERPLUS_EXALLC | IVW | 14 | -0.103 | 0.022 | $1.83 \times 10^{-6}$ | 0.902 (0.865 - 0.941) |

IVW, Inverse variance weighted.

For breast cancer subtypes, our research revealed that N-acetylglutamate is casually associated with an increased risk of HER-breast cancer (OR = 1.429, 95% CI = 1.198 - 1.706, p = 7.51×10^−5^, IVW), whereas indolepropionate is causally associated with a reduced risk of HER-breast cancer (OR = 0.746, 95% CI = 0.647 - 0.860, p = 5.03×10^−5^, IVW). This study also revealed that DHEAS (OR = 1.294, 95% CI = 1.175 - 1.424, p = 1.48×10^−7^, IVW) and androsterone sulfate (OR = 1.071, 95% CI = 1.039 - 1.105, p = 1.01×10^−5^, IVW) are causally associated with an increased risk of HER+ breast cancer. Ribitol (OR = 0.894, 95% CI = 0.854 - 0.936, p = 1.42×10^−6^, IVW), homostachydrine (OR = 0.581, 95% CI = 0.478 - 0.705, p = 4.23×10^−8^, IVW), and glyco-beta-muricholate (OR = 0.902, 95% CI = 0.865 - 0.941, p = 1.83×10^−6^, IVW) are causally associated with a reduced risk of HER+ breast cancer.

### Causal effects independent of cancer-related factors

Prior research has established associations between lifestyle factors and cancer risks. These lifestyle factors include smoking^9^, alcohol consumption^10^, BMI ^11^, physical exercise ^12^, and chemical exposure ^13^. To assess the influence of these covariates on causal relationships, we conducted an MVMR analysis adjusting to these covariates. Among the 694 significant causal associations (p < 1×10^−5^), 374 causal associations were validated by the MVMR analysis. Notably, the MVMR analyses revealed similar causal effects of BTN2A1 and BTN3A2 on cancer risks as those identified by UVMR analysis (Figure 2). The MVMR results are presented in our online database.

**Figure 2.**
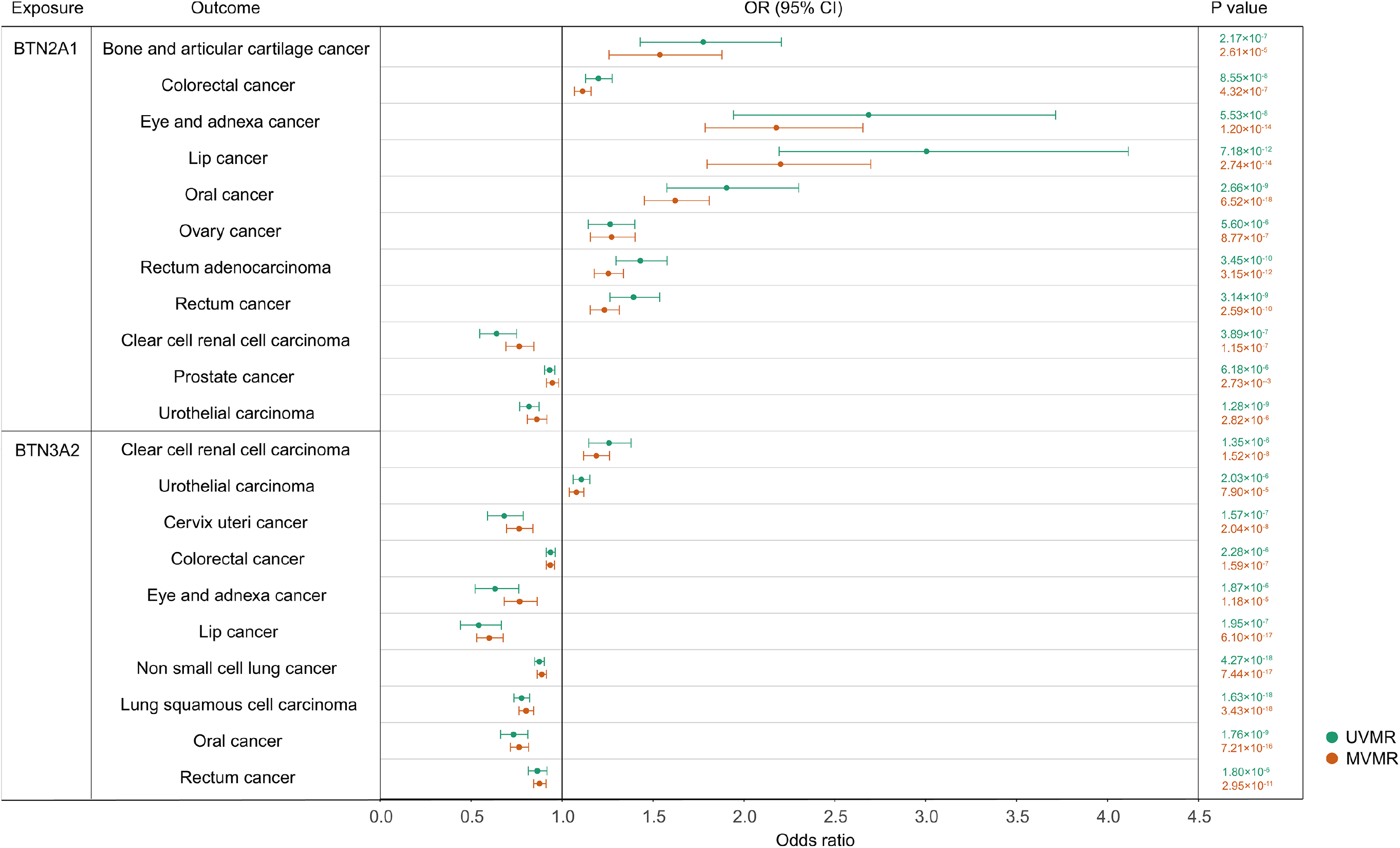
MVMR analyses of the causal effect of BTN2A1 and BTN3A2 on cancer risks. MVMR analyses demonstrated similar causal efforts of BTN2A1 and BTN3A2 as those identified by UVMR analyses.

## Discussion

The investigation of causal relationships is critical for elucidating the driving factors behind observed associations. In this study, we employed MR analyses to identify the causal effects of plasma proteins and metabolites on cancers. To the best of our knowledge, this study, encompassing 2,714 proteins, 731 metabolite levels, 235 metabolite ratios, and 99 types of cancers, is the most comprehensive analysis to date on the causal effects of plasma proteins and metabolites on cancer risks.

Cancer risk assessment is significant for cancer prevention and understanding cancer mechanisms. Advances in proteomics and metabolomics technologies have fueled interest in exploring the clinical utility of circulating proteins and metabolites as non-invasive biomarkers, given their ability to reflect physiological processes. Although prior studies have identified plasma proteins and metabolites involved in cancer mechanisms, their roles in risk assessment and prevention were constrained by unclear causal associations. Benefiting from recent large-scale GWAS analyses and the methodology of MR analysis, we systematically investigated the potential causal relationships between plasma proteins and metabolites and the risk of various cancers. Our findings significantly contribute to cancer risk prediction and the identification of new therapeutic targets, thereby improving cancer management and treatment strategies.

Gamma delta (γδ) T cells are essential to anti-tumor immunity. BTN3A1 and BTN2A1 are crucial for Vγ9Vδ2 T cell response to phosphoantigens (pAgs) overexpressed in tumor cells ^20,21 22 23^. Previous studies revealed that BTN2A1 is correlated with advanced renal cell carcinoma ^24^, ovarian cancer ^25^, breast cancer ^26^, and head and neck squamous cell carcinoma ^27^. Our study demonstrated that BTN2A1 is causally associated with an increased risk of multiple cancer types, including colorectal cancer, oral cancer, ovary cancer, rectal cancer, and others. This result indicates the role of Vγ9Vδ2 T cells in these cancer types. However, we observed that BTN2A1 is causally linked to a reduced risk of several urologic neoplasms such as ccRCC, prostate cancer, and urothelial carcinoma, suggesting that Vγ9Vδ2 T cells may function differently in these urologic neoplasms. On the other hand, BTN3A2, another member of the butyrophilins, is not as well studied as BTN2A1 and BTN3A1. Our investigation revealed that BTN3A2 is causally associated with reduced risks of multiple cancer types, indicating its protective role in cancer development.

This investigation also identified some metabolites causally associated with cancer risks. N6-carbamoylthreonyladenosine is correlated with chronic kidney disease ^28^, hypertension ^29^, and hepatorenal dysfunction ^30^. Previous studies have revealed that N6-carbamoylthreonyladenosine is causally related to an increased risk of colorectal and lung cancer and a reduced risk of breast cancer ^31^. Beyond these results, our study demonstrated a causal association between N6-carbamoylthreonyladenosine and basal cell carcinoma, cervix uteri cancer, colorectal cancer, diffuse large B cell lymphoma, eye and adnexa cancer, head and neck cancer, and hepatocellular carcinoma. Our investigation has extended the causal effect of N6-carbamoylthreonyladenosine to a broader range of cancer types and may inspire the mechanisms of N6-carbamoylthreonyladenosine in tumorigenesis.

Nevertheless, this study has several limitations. First, our findings are derived from a European population, potentially leading to biased estimates and limited generalizability. Future research is required to investigate these causal relationships in diverse populations. Second, the strict criteria for multiple-testing correction may have resulted in the omission of some causal associations. Third, the F-statistic for some exposure data is below 10, warranting caution regarding weak IV bias. Fourth, experimental studies are still necessary to elucidate the mechanisms through which plasma proteins and metabolites affect cancer development.

## Conclusion

In summary, this study provides a comprehensive assessment of the causal effect of plasma proteins and metabolites on various cancers using two-sample MR analysis. We established causal associations between plasma proteins and cancers as well as causal associations between plasma metabolites and cancers. Additionally, we built an online database containing all results, which may be enlightening for other scientists. This study not only sheds light on the mechanisms of cancer development but also holds significant implications for the prevention and targeted therapy of cancers.

## Data availability

The plasma protein GWAS summary statistics were downloaded from the UKB-PPP website (https://www.synapse.org/%23!Synapse:syn51365301). The plasma metabolite GWAS summary statistics were downloaded from the GWAS Catalog website (https://www.ebi.ac.uk/gwas/). The GWAS summary statistics from FinnGen were downloaded following the instruction on its website (https://www.finngen.fi/en/access_results).

## Code availability

No special codes were used in this study. The codes used in MR analysis with two-sample MR R package are available on the GitHub repository (https://github.com/BowenDuGroup/CAURIS).

## Acknowledgements

The data analyzed in this study was provided by UKB-PPP, plasma metabolome, and FinnGen projects. We gratefully acknowledge their contributing studies and the participants in these studies.

## Authors’ contributions

B.D. and X.S. designed the study. B.D. analyzed the data. B.D. L.F., and C.T. curated and processed the data. B.D. wrote the manuscript. J.G and X.S. supervised the research. All authors read and approved the final manuscript.

## Ethics statement

This study is performed using published studies and publicly available summary statistics. All original studies have been approved by the corresponding ethical review board. The participants have provided informed consent. Therefore, no new ethical review board approval was required.

## Reference

1. Siegel RL, Miller KD, Wagle NS, Jemal A. Cancer statistics, 2023. CA Cancer J Clin 2023; 73(1): 17–48.

2. Davies MPA, Sato T, Ashoor H, et al. Plasma protein biomarkers for early prediction of lung cancer. EBioMedicine 2023; 93: 104686.

3. Kurki MI, Karjalainen J, Palta P, et al. FinnGen provides genetic insights from a well-phenotyped isolated population. Nature 2023; 613(7944): 508–18.

4. Long Y, Tang L, Zhou Y, Zhao S, Zhu H. Causal relationship between gut microbiota and cancers: a two-sample Mendelian randomisation study. BMC Med 2023; 21(1): 66.

5. Burgess S, Thompson SG, Collaboration CCG. Avoiding bias from weak instruments in Mendelian randomization studies. Int J Epidemiol 2011; 40(3): 755–64.

6. Burgess S, Davies NM, Thompson SG. Bias due to participant overlap in two-sample Mendelian randomization. Genet Epidemiol 2016; 40(7): 597–608.

7. Bowden J, Davey Smith G, Burgess S. Mendelian randomization with invalid instruments: effect estimation and bias detection through Egger regression. Int J Epidemiol 2015; 44(2): 512–25.

8. O’Connor LJ, Price AL. Distinguishing genetic correlation from causation across 52 diseases and complex traits. Nat Genet 2018; 50(12): 1728–34.

9. Hecht SS, Hatsukami DK. Smokeless tobacco and cigarette smoking: chemical mechanisms and cancer prevention. Nat Rev Cancer 2022; 22(3): 143–55.

10. Rumgay H, Murphy N, Ferrari P, Soerjomataram I. Alcohol and Cancer: Epidemiology and Biological Mechanisms. Nutrients 2021; 13(9).

11. Pati S, Irfan W, Jameel A, Ahmed S, Shahid RK. Obesity and Cancer: A Current Overview of Epidemiology, Pathogenesis, Outcomes, and Management. Cancers (Basel) 2023; 15(2).

12. Rodriguez-Canamero S, Cobo-Cuenca AI, Carmona-Torres JM, et al. Impact of physical exercise in advanced-stage cancer patients: Systematic review and meta-analysis. Cancer Med 2022; 11(19): 3714–27.

13. Koual M, Tomkiewicz C, Cano-Sancho G, Antignac JP, Bats AS, Coumoul X. Environmental chemicals, breast cancer progression and drug resistance. Environ Health 2020; 19(1): 117.

14. Li W, Chen R, Feng L, et al. Genome-wide meta-analysis, functional genomics and integrative analyses implicate new risk genes and therapeutic targets for anxiety disorders. Nat Hum Behav 2024; 8(2): 361–79.

15. Song Z, Li S, Shang Z, et al. Integrating multi-omics data to analyze the potential pathogenic mechanism of CTSH gene involved in type 1 diabetes in the exocrine pancreas. Brief Funct Genomics 2023.

16. Peris K, Fargnoli MC, Kaufmann R, et al. European consensus-based interdisciplinary guideline for diagnosis and treatment of basal cell carcinoma-update 2023. Eur J Cancer 2023; 192: 113254.

17. His M, Viallon V, Dossus L, et al. Prospective analysis of circulating metabolites and breast cancer in EPIC. BMC Med 2019; 17(1): 178.

18. Zhao H, Shen J, Ye Y, et al. Validation of plasma metabolites associated with breast cancer risk among Mexican Americans. Cancer Epidemiol 2020; 69: 101826.

19. Brantley KD, Zeleznik OA, Rosner B, et al. Plasma Metabolomics and Breast Cancer Risk over 20 Years of Follow-up among Postmenopausal Women in the Nurses’ Health Study. Cancer Epidemiol Biomarkers Prev 2022; 31(4): 839–50.

20. Rigau M, Ostrouska S, Fulford TS, et al. Butyrophilin 2A1 is essential for phosphoantigen reactivity by gammadelta T cells. Science 2020; 367(6478).

21. Yuan L, Ma X, Yang Y, et al. Phosphoantigens glue butyrophilin 3A1 and 2A1 to activate Vgamma9Vdelta2 T cells. Nature 2023; 621(7980): 840–8.

22. Willcox CR, Salim M, Begley CR, et al. Phosphoantigen sensing combines TCR-dependent recognition of the BTN3A IgV domain and germline interaction with BTN2A1. Cell Rep 2023; 42(4): 112321.

23. Cano CE, Pasero C, De Gassart A, et al. BTN2A1, an immune checkpoint targeting Vgamma9Vdelta2 T cell cytotoxicity against malignant cells. Cell Rep 2021; 36(2): 109359.

24. Billon E, Chanez B, Rochigneux P, et al. Soluble BTN2A1 Is a Potential Prognosis Biomarker in Pre-Treated Advanced Renal Cell Carcinoma. Front Immunol 2021; 12: 670827.

25. Fanale D, Corsini LR, Brando C, et al. Can circulating PD-1, PD-L1, BTN3A1, pan-BTN3As, BTN2A1 and BTLA levels enhance prognostic power of CA125 in patients with advanced high-grade serous ovarian cancer? Front Oncol 2022; 12: 946319.

26. Ren H, Li S, Liu X, Li W, Hao J, Zhao N. Multi-omics analysis of the expression and prognostic value of the butyrophilins in breast cancer. J Leukoc Biol 2021; 110(6): 1181–95.

27. Lu H, Dai W, Guo J, et al. High Abundance of Intratumoral gammadelta T Cells Favors a Better Prognosis in Head and Neck Squamous Cell Carcinoma: A Bioinformatic Analysis. Front Immunol 2020; 11: 573920.

28. Denburg MR, Xu Y, Abraham AG, et al. Metabolite Biomarkers of CKD Progression in Children. Clin J Am Soc Nephrol 2021; 16(8): 1178–89.

29. Al Ashmar S, Anwardeen NR, Anlar GG, Pedersen S, Elrayess MA, Zeidan A. Metabolomic profiling reveals key metabolites associated with hypertension progression. Front Cardiovasc Med 2024; 11: 1284114.

30. Mindikoglu AL, Opekun AR, Putluri N, et al. Unique metabolomic signature associated with hepatorenal dysfunction and mortality in cirrhosis. Transl Res 2018; 195: 25–47.

31. Chen Y, Xie Y, Ci H, et al. Plasma metabolites and risk of seven cancers: a two-sample Mendelian randomization study among European descendants. BMC Med 2024; 22(1): 90.

